## Supplementary material for "Comparison of semaglutide in combined with empagliflozin versus semaglutide and empagliflozin monotherapy in non-alcoholic fatty liver disease with type 2 diabetes: study protocol for a randomised controlled clinical trial": Informed consent form.docx

**INFORMED CONSENT FORM-** INFORMED PAGE

**Trial Name**: Comparison of efficacy and safety of glucagon-like peptide-1 receptor agonists, sodium-glucose cotransporter 2 inhibitors, and their combination in patients with type 2 diabetes and non-alcoholic fatty liver disease

**Principal investigator**: Jian-Qing Tian

**Sponsor**: Jian-Qing Tian

Dear Subjects:

You are invited to participate in a "clinical trial". Please read this informed consent carefully and decide whether to participate in it. When the doctor or researcher discusses the informed consent form with you, feel free to ask for clarification on anything that is unclear. We recommend discussing it with your family and friends before joining this clinical trial. If you are already involved in another study, please inform the doctor or researcher accordingly. The objectives, procedures, and other essential information regarding this clinical trial are outlined below.

**1. Objective of trails:**

**Main Objectives:** Comparison of efficacy and safety of glucagon-like peptide-1 receptor agonists, sodium-glucose cotransporter 2 inhibitors, and their combination in patients with type 2 diabetes and non-alcoholic fatty liver disease.

**Secondary Objectives:** To investigate the mechanism of glucagon-like peptide-1 receptor agonists and sodium-glucose cotransporter 2 inhibitors in patients with type 2 diabetes mellitus and non-alcoholic fatty liver disease.

**2. Process of trials:**

**How many people will participate in this trail?**

Approximately 105 people will participate in this trail at Xiamen Humanity Hospital, Fujian Medical University.

**Procedure of trails**

Population screening was conducted based on inclusion and exclusion criteria, and individuals who met the criteria were included in the study. The included participants were randomly assigned to receive either glucagon-like peptide-1 receptor agonists, sodium-glucose cotransporter 2 inhibitors and glucagon-like peptide-1 receptor agonists plus sodium-glucose cotransporter 2 inhibitors.

**You will be eligible for the trial if you meet the following inclusion criteria:**

1. Age of ≥18 years old at the time of enrolment.

2. Diagnosis of non-alcoholic fatty liver disease.

3. Diagnosis of type 2 diabetes.

4. Body mass index (BMI) more than 24 kg/m2.

5. Patients who had not used glucagon-like peptide-1 receptor agonist and sodium-glucose cotransporter 2 inhibitors within 4 weeks.

**You will not be eligible for the trial if you meet the following exclusion criteria:**

1. Type 1 diabetes mellitus, gestational diabetes mellitus or other types of diabetes mellitus.

2. Previously treated with thiazolidinediones.

3. History of cardiac, hepatic or renal insufficiency.

4. Patients with viral hepatitis (such as hepatitis B), alcoholic liver disease, autoimmune hepatitis, drug-induced liver disease, hemochromatosis, Wilson’s disease, liver cirrhosis, inborn errors of metabolism (such as cholesterol ester storage disease), or other causes of chronic liver disease.

5. History of cerebral stroke, malignant tumor or pancreatitis.

6. History of medullary thyroid carcinoma or multiple endocrine neoplasia type 2 in oneself or family.

7. Pregnancy, lactation or desire for conception during the study period.

8. Consumed more than 140g of ethanol per week for men and more than 70g of ethanol per week for women.

9. Known or suspected hypersensitivity to GLP-1 receptor agonists, SGLT-2 inhibitors or metformin.

10. Known or suspected mental and psychological disorders.

11. History of chronic anaemia (Hb level <100 g/L in men and <90 g/L in women).

12. Unwilling or unable to provide informed consent.

**3. Study flow**

Before enrolling in a study, you will undergo the following tests during the screening period to determine your eligibility for participation:

Transient elastography (FibroScan).

If you qualify for the above tests, you will participate in the following steps, known as the treatment period (or visit period):

**Treatment:**

1. glucagon-like peptide-1 receptor agonist,

2. sodium-glucose cotransporter 2 inhibitors,

3. glucagon-like peptide-1 receptor agonist plus sodium-glucose cotransporter 2 inhibitors.

**Follow-up:**

1. transient elastography (FibroScan) will be conducted at baseline and, weeks 24, 36 and 52.

2. free fatty acid, glucagon, Routine biochemical, routine blood, glycated hemoglobin, fasting insulin, fasting C-peptide, bone metabolism, interleukin-6, adiponectin, high-sensitivity CRP and ferritin will be measured at baseline and weeks 12, 24, 36 and 52.

3. anthropometric indicators will be measured at baseline and weeks 12, 24, 36 and 52.

4. the symptoms of gastrointestinal discomfort and reproductive tract infection were observed in real time.

After completing your treatment, you will also continue to receive end-of-treatment visits and long-term follow-up (or survival follow-up) in order to monitor for any adverse reactions and subsequent treatment.

**End of treatment/Withdraw the study:**

When subjects end the study treatment or withdraw from the study, certain assessments and examinations will be conducted as listed：

Survival follow-up,

before disease progression visit,

after disease progression visit,

how long will the study last?

**4. Risk and benefit**

**What are the risks of participating in this study:**

Specific adverse events: abdominal pain, nausea, vomiting、urinary frequency, urinary urgency, urinary painful.

**What are the benefits of participating in the study:**

Benefited in direct way: participants were paid for completing the trial.

Benefit from potentially way: physician consultations are free for all included participants; Glycemic control will be improved.

**5. Use of research results and confidentiality of personal information**

Personal information for both potential and enrolled participants will be kept confidential before, during and after the trial. Participants personal information will not be included in any study forms, reports, publications or any other disclosures unless legally required.

**6. Participants’** **Rights and responsibilities**

**Rights**

You are free to decide whether or not to participant in this research study. If you decide to participant, you will receive a copy of this information sheet and your signed consent form. If you choose not to participant, it will not affect any other treatments you should receive.

**Responsibilities**

The symptoms of discomfort should be reported to the doctor in real time.

**7. Contact information:**

If you have any questions related to this study, please contact Yu-Hao Lin at 15868322740. If you have any questions regarding your rights or if you would like to report any difficulties, grievances, or concerns encountered during your participation in this study, or if you would like to provide comments and suggestions related to this study, please contact the Ethics Committee at the following: 0592-5261060 or email them at.

*（Informed notification completed, the following is the informed consent signature page）*

**INFORMED CONSENT FORM-** SIGNATURE PAGE

**Subject name:**

**Contact number:**

**【Subject confirmation】：**

I have been informed of the purpose, background, process, risks and benefits of the study. I had ample time and opportunity to ask questions, and I was content with the answers.

I was also given information on whom to contact in case of questions, difficulties, concerns, suggestions for research or if I needed further information or help with my research.

I have read this informed consent form and agree to participate in this study.

**I am aware that I have the option to decline participation or withdraw from the study at any time without providing a reason.**

I am aware that if my condition worsens, or if I experience serious adverse events, or if my study doctor determines that it is not in my best interests to continue participating in the study, he or she will advise me to withdraw. The sponsor or regulatory authority may also terminate the study during the study period without obtaining my consent. In such a case, the doctor will promptly inform me and provide alternative treatments.

**I will get a copy of this informed consent with my signature and that of the investigator.**

**Subject signature:** **Date:**

(Note: If the subject does not have full capacity for civil conduct, the legal representative shall sign at the signature of the legal representative below)

**Signature of legal representative:**  **Date:**

**Contact number:**

**Investigator's statement:** I have accurately informed the subject of this document, that he/she read the informed consent form accurately, and I certify that the subject was given an opportunity to ask questions and voluntarily consented.

**Investigator signature:**	**Date**:

**Contact number:**
