## Supplementary material for "Comparison of semaglutide in combined with empagliflozin versus semaglutide and empagliflozin monotherapy in non-alcoholic fatty liver disease with type 2 diabetes: study protocol for a randomised controlled clinical trial": Research Protocol.docx

**Principle Investigator:** Yu-Hao Lin

**Organizer:** Xiamen Humanity Hospital, Fujian Medical University, Xiamen, Fujian, China.

### 1. Integrity statement

Our research group solemnly declares that the process of this study is carried out in accordance with the study plan, and the study data are truly and accurately recorded. The relevant research results, including technical standards, patents and other related intellectual property rights belong to the research group. The research group is fully aware of the legal responsibilities to be borne in this statement. There is no conflict of interest in this research group.

### 2. Funding

Medical and Health Guidance Projects of Xiamen, China (3502Z20224ZD1106)

### 3. Background

Diabetes is a metabolic disease characterized by chronic hyperglycemia, which is caused by deficiencies in insulin secretion and/or utilization. Nonalcoholic fatty liver disease (NAFLD) is a liver disease that is associated with obesity and metabolic dysfunction, and it is closely related to diabetes. Epidemiological surveys in East Asian populations have shown that the prevalence of NAFLD in people with diabetes is 55%, which seriously harm human health and imposes a huge economic burden on society. ^(1)^

Type 2 diabetes mellitus (T2DM) often coexists with NAFLD, and T2DM is an important factor to accelerate the progression of NAFLD. T2DM reduces the sensitivity of liver tissue to insulin action, leading to insulin resistance. Insulin resistance is considered to be the central link in the pathogenesis of NAFLD. ^(2)^ Insulin resistance can drive the de novo synthesis of fat in the liver, resulting in an increase in the level of free fatty acids, thereby increasing the utilization of free fatty acids, resulting in the production of excess adipocytes in the liver and pancreas and the formation of fatty liver and pancreas.^(3)^ Fatty liver and pancreas can lead to β cell dysfunction and insulin resistance through a variety of ways, forming a negative cycle. Therefore, it is speculated that the disease progression can be controlled by stabilizing the glucose and lipid metabolism levels in patients with T2DM complicated with NAFLD.

At present, there is no specific drug for the treatment of NAFLD. Glucagon-like peptide-1 (GLP-1) receptor agonists and sodium-glucose cotransporter 2 (SGLT-2) inhibitors, as antidiabetic drugs, have the effect of reducing body weight and improving liver steatosis. ^(4)^ Further, a real-world study has found that GLP-1 receptor agonists and SGLT-2 inhibitors can significantly improve liver steatosis and fibrosis in patients with NAFLD with T2DM. ^(5)^ Kuchay et al. have found that dulaglutide, a GLP-1 receptor agonists, can significantly reduce the intrahepatic fat content compared with control group in T2DM patients with NAFLD, its effects were no significant correlation with body weight and glucose reduction. ^(6)^ In another study, compared with control group, semaglutide, as a GLP-1 receptor agonist, improved hepatic steatosis and glucose levels in patients with nonalcoholic steatohepatitis. ^(7)^ Another study found that the empagliflozin, as a SGLT-2 inhibitor can significantly reduce intrahepatic fat content and blood glucose levels in patients with T2DM and NAFLD. And its effect was not significantly correlated with reduction in blood glucose or body weight. ^(8)^

At present, there have been few studies on the effects of combination of GLP-1 receptor agonists and SGLT-2 inhibitors in patients with NAFLD. Gastaldelli et al. have found that combination of GLP-1 receptor agonists and SGLT-2 inhibitors can improve the liver related biological indicators and liver steatosis in patients with T2DM. ^(9)^ We hypothesized that the combination of semaglutide and empagliflozin can improve hepatic steatosis and glycemic control in patients with T2DM and NAFLD.

Although current studies have shown that GLP-1 receptor agonists and SGLT-2 inhibitors can reduce intrahepatic fat content and delay disease progression in patients with NAFLD. However, its mechanism has not been fully elucidated. Some studies have found that glucagon can induce amino acid catabolism, promoting hepatic β-oxidation and insulin secretion, reducing lipogenesis and the concentration of circulating free fatty acids, thereby reducing the accumulation of fat in the liver and pancreas. ^(10, 11)^ NAFLD can impair the catabolism of amino acids induced by glucagon. Therefore, we speculate that semaglutide and empagliflozin can delay the progression of NAFLD by the effect of glucagon on amino acids and free fatty acids.

### 4. Objectives

This study planned to carry out a clinical trial in which patients with type 2 diabetes mellitus and non-alcoholic fatty liver disease were randomly divided into three groups and given semaglutide, empagliflozin, and semaglutide plus empagliflozin, respectively. The effects of each group on intrahepatic fat content, diabetes remission rate, and body weight were studied. The incidence of gastrointestinal symptoms and genital tract infection were used to evaluate the safety differences among the three groups. Glucagon and free fatty acids were further measured to explore the mechanism of semaglutide and empagliflozin in non-alcoholic fatty liver disease, in order to provide theoretical basis for clinical application.

### 5. Inclusion and exclusion criteria

**Inclusion criteria**

1. Age of ≥18 years old at the time of enrolment.

2. Diagnosis of NAFLD.

3. Diagnosis of T2DM.

4. Body mass index (BMI) more than 24 kg/m^2^.

5. Patients who had not used GLP-1 receptor agonists and SGLT-2 inhibitors within 4 weeks.

**Exclusion criteria**

1. Type 1 diabetes mellitus, gestational diabetes mellitus or other types of diabetes mellitus.

2. Previously treated with thiazolidinediones.

3. History of cardiac, hepatic or renal insufficiency.

4. Patients with viral hepatitis (such as hepatitis B), alcoholic liver disease, autoimmune hepatitis, drug-induced liver disease, hemochromatosis, Wilson’s disease, liver cirrhosis, inborn errors of metabolism (such as cholesterol ester storage disease), or other causes of chronic liver disease.

5. History of cerebral stroke, malignant tumor or pancreatitis.

6. History of medullary thyroid carcinoma or multiple endocrine neoplasia type 2 in oneself or family.

7. Pregnancy, lactation or desire for conception during the study period.

8. Consumed more than 140g of ethanol per week for men and more than 70g of ethanol per week for women.

9. Known or suspected hypersensitivity to GLP-1 receptor agonists, SGLT-2 inhibitors or metformin.

10. Known or suspected mental and psychological disorders.

11. History of chronic anaemia (Hb level <100 g/L in men and <90 g/L in women).

12. Unwilling or unable to provide informed consent.

### 6. Design scheme

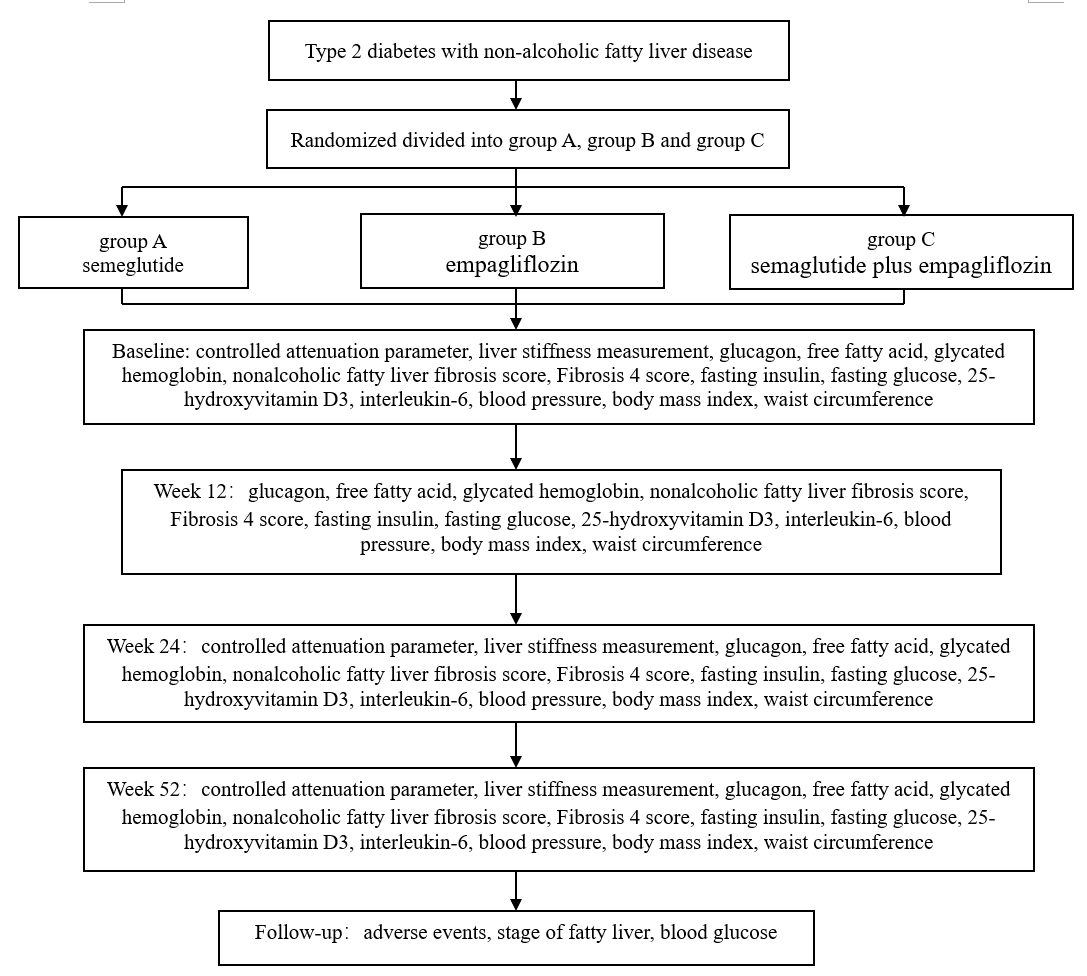
A total of 105 patients with type 2 diabetes mellitus and non-alcoholic fatty liver disease were enrolled in this 52-weeks prospective randomized controlled trail. According to the inclusion and exclusion criteria, the enrolled patients were randomly divided into semaglutide group (group A), empagliflozin group (group B), and semaglutide plus empagliflozin group (group C). The outcomes will evaluate on the day of enrollment, weeks 12, 24 and 52. The detailed technical roadmap is as follows:

### 7. Sample Size Estimation:

With α=0.05 and Power=0.90, the sample size was calculated by One-way Analysis of Variance (ANOVA) based on means in NCSS-PASS. Each group needed (8+27+58)/3=31. The dropout rate was estimated at 10%, which required 31/ (1-0.1) =35 cases in each group, and 35*3=105 cases in total.

### 8. Randomization and concealed

According to the centre by statistical grouping computer random number. Random numbers were concealed in airtight brown paper envelopes.

### 9. Blinding

Blinding of researchers and patients.

### 10. Outcomes

(1) Primary outcomes:

1. Degree of fatty liver was measured by liver ultrasound at baseline and, 24 and 52 weeks.

2. Glucagon was measured by ELISA at baseline and, 12, 24 and 52 weeks.

3. free fatty acids was measured by ELISA at baseline and, 12, 24 and 52 weeks.

(2) Secondary outcomes:

1. Liver stiffness was measured by liver ultrasound at baseline and, 24 and 52 weeks.

2. Nonalcoholic fatty liver fibrosis score was calculated by the formula at baseline and, 12, 24 and 52weeks.

3. Fibrosis-4 index was calculated by the formula was at baseline and, 12, 24 and 52 weeks.

4. Fasting insulin was measured by the direct chemiluminescence at baseline and, 12, 24 and 52 weeks.

5. Fasting blood glucose was measured by enzyme chemical at baseline and, 12, 24 and 52 weeks.

6.25-hydroxyvitamin D3 was measured by enzyme chemical at baseline and, 12, 24 and 52 weeks.

7. Interleukin-6 was measured by ELISA at baseline and, 12, 24, 52 weeks.

8. Ferritin was measured by chemiluminescence immunoassay at baseline and, 12, 24 and 52 weeks.

9. Blood pressure was measured by sphygmomanometer at baseline and, 12, 24 and 52 weeks.

10.Body mass index was calculated by formula at baseline and, 12, 24 and 52 weeks.

11. Adiponectin was measured by ELISA at baseline and, 12, 24 and 52 weeks.

(3) Safety outcomes:

The patients' symptoms of gastrointestinal discomfort and genital tract infection were observed in real time. The number of adverse events was counted at 12, 24 and 52 weeks, and the rate was calculated.

### 11. The definition of the validity of the study participants

1. The subject has the right to withdraw from the trial at any stage of the trial.

2. Withdrawal from the trial if the following conditions occur during the trial:

I. Intolerance of glucagon-like peptide-1 receptor agonists or sodium-glucose cotransporter 2 inhibitors during the trial.

II. Patients with acute complications of diabetes (such as diabetic ketoacidosis, diabetic hyperosmolar hyperglycemia, etc.) during the period.

III. Patients who failed to use semaglutide and empagliflozin on time.

IV. Patients who fail to follow up on time.

3. The investigators informed the subjects of their contact information and obtained the latest contact information of the subjects actively.

### 12. Definition, identification and management of adverse events

1. Adverse event is defined as any medical adverse events that occurs in a patient or clinical research subject while receiving a research intervention, whether it is related to the intervention or not.

2. Adverse events were defined as the occurrence of pancreatitis, gastrointestinal reactions such as nausea, vomiting, and urinary tract infection.

3. When adverse events occur, symptomatic treatment and discontinuation of the drug are used based to the patient's tolerance and the severity of adverse events.

### 13. Recruitment of participants

Recruitment site is Xiamen Humanity Hospital, Fujian Medical University, Xiamen.

Notices about recruitment are displayed on bulletin boards within the hospital, as well as on WeChat friend circle, WeChat group, Weibo and other social media platforms. The eligibility of potential participants will be evaluated by the study coordinator. All participants will be required to sign an informed consent form if they agree to participate in the study.

### 14. Participants generally information collection

The endocrinologists of Xiamen Humanity Hospital, Fujian Medical University collected the participants' mobile phone number, WeChat, home address, and contact information of the main contacts.

### 15. Statistical analysis

1. T-test was used to verify the difference between the two groups of data for quantitative data conforming to normal distribution.

2. Analysis of variance was used to verify the difference of multiple groups of data for quantitative data conforming to normal distribution.

3. Chi-square test was used for qualitative data to verify the difference between groups.

4. Kruskal-Wallis test was used to verify the difference between the data of different groups for the quantitative data that did not conform to the normal distribution.

5. Logistics regression analysis was used to analyze the correlation between variables.

When the number of people decreased for various reasons during the trial, the outcomes were analyzed by intention to treat.

### 16. Management system

Participants join WeChat group at the time of enrollment and are managed by professionals.

Test specimens were collected by professional nurses and tested and save by clinical laboratory doctors.

Data was collected and recorded by professionals, stored in Excel spreadsheet, and checked by another professional.

### 17. Treatment and management of participants after the trial

If participants still fail to improve fatty liver and blood glucose after the trial, treatments will be changed, including but not limited to increasing the use of hypoglycemic drugs and insulin.
